# The COVID-19 pandemic and its prolonged impacts on food prices, food consumption and diet quality in sub-Saharan Africa

**DOI:** 10.1101/2022.12.12.22283393

**Authors:** Abbas Ismail, Isabel Madzorera, Edward A. Apraku, Amani Tinkasimile, Dielbeogo Dasmane, Pascal Zabre, Millogo Ourohire, Nega Assefa, Angela Chukwu, Firehiwot Workneh, Frank Mapendo, Bruno Lankoande, Elena Hemler, Dongqing Wang, Sulemana W. Abubakari, Kwaku P. Asante, Till Baernighausen, Japhet Killewo, Ayoade Oduola, Ali Sie, Abdramane Soura, Said Vuai, Emily Smith, Yemane Berhane, Wafaie W. Fawzi

**Affiliations:** College of Natural and Mathematical Sciences, University of Dodoma, Dodoma, Tanzania; Division of Community Health Sciences, School of Public Health, University of California, Berkeley, 2121 Berkeley Way, Berkeley, CA, 94720, United States of America; Kintampo Health Research Center, Research and Development Division, Ghana Health Service, Box 200, Kintampo, Bono East Region, Ghana; Africa Academy for Public Health, Dar es Salaam, Tanzania; Institut Supérieur des Sciences de la Population, University of Ouagadougou, Ouagadougou, Burkina Faso; Nouna Health Research Center, Burkina Faso; College of Health and Medical Sciences, Haramaya University, Harar, Ethiopia; Department of Statistics, University of Ibadan, Ibadan, Nigeria; Addis Continental Institute of Public Health, Ethiopia; Department of Global Health and Population, Harvard T.H. Chan School of Public Health, Harvard University, Boston, Massachusetts, United States of America; Heidelberg Institute of Global Health, University of Heidelberg, Heidelberg, Germany; Department of Epidemiology and Biostatistics, Muhimbili University of Health and Allied Sciences, Dar es Salaam, Tanzania; University of Ibadan Research Foundation, University of Ibadan, Ibadan, Nigeria; Department of Global Health, Milken Institute School of Public Health, George Washington University, Washington, DC, United States of America; Department of Exercise and Nutrition Sciences, Milken Institute School of Public Health, George Washington University, Washington, DC, United States of America; Department of Nutrition, Harvard T.H. Chan School of Public Health, Harvard University, Boston, Massachusetts, United States of America; Department of Epidemiology, Harvard T.H. Chan School of Public Health, Harvard University, Boston, Massachusetts, United States of America

**Keywords:** COVID-19, Diet quality, Prime Diet Quality Score, food prices, food consumption, food security, Minimum Dietary Diversity for Women, Sub-Saharan Africa

## Abstract

**Background:** Sub-Saharan Africa faces prolonged COVID-19 related impacts on economic activity, livelihoods, nutrition, and food security, with recovery slowed down by lagging vaccination progress.

**Objective:** This study investigated the economic impacts of COVID-19 on food prices, consumption and dietary quality in Burkina Faso, Ethiopia, Ghana, Nigeria, and Tanzania.

**Methods:** We conducted a repeated cross-sectional study and used a mobile platform to collect data. Data collected from round 1 (July-November, 2020) and round 2 (July-December, 2021) were considered. We assessed participants’ dietary intake of 20 food groups over the previous seven days. The study’s primary outcome was the Prime Diet Quality Score (PDQS), with higher scores indicating better dietary quality. We used linear regression and generalized estimating equations to assess factors associated with diet quality during COVID-19.

**Results:** Most of the respondents were male and the mean age (±SD) was 42.4 (±12.5) years. Mean PDQS (±SD) was low at 19.1 (±3.8) before COVID-19, 18.6(±3.4) in Round 1, and 19.4(±3.8) in Round 2. A majority of respondents (80%) reported higher than expected prices for all food groups during the pandemic. Secondary education or higher (estimate: 0.73, 95% CI: 0.32, 1.15), older age (estimate: 30-39 years: 0.77, 95% CI: 0.35, 1.19, or 40 years or older: 0.72, 95% CI: 0.30, 1.13), and medium wealth status (estimate: 0.48, 95% CI: 0.14, 0.81) were associated with higher PDQS. Farmers and casual laborers (estimate: -0.60, 95% CI: -1.11, - 0.09), lower crop production (estimate: -0.87, 95% CI: -1.28, -0.46) and not engaged in farming (estimate: -1.38, 95% CI: -1.74, -1.02) associated with lower PDQS.

**Conclusion:** Diet quality which had declined early in the pandemic had started to improve. However, consumption of healthy diets remained low, and food prices remained high. Efforts should continue to improve diet quality for sustained nutrition recovery through mitigation measures, including social protection.

## Introduction

The coronavirus disease 2019 (COVID-19) continues to affect the social, economic and health status of individuals and communities globally and has exposed significant inequalities by income, socio-demographic factors and geographic locations (1). Despite the persistent improvement of global health, some settings continue to face greater threats to health and well-being during public health emergencies due to prevailing social, economic, political, and environmental conditions, and the COVID-19 pandemic is not an exception with greater impacts on the socially disadvantaged, including on the African continent (2, 3).

The absolute number of reported cases and mortality due to COVID-19 in Africa has been lower than in other regions, with 8.4 million cases and 170,300 deaths reported by March 2022 (4). However, challenges with emerging variants are likely to continue in the region due to vaccine hesitancy and low vaccination coverage (5), in contrast to declining global cases and recovery of economies (5, 6). Further, Sub-Saharan Africa (SSA) was already grappling with economic and health challenges before the pandemic, so the impacts of COVID-19 could be more long-lasting than in developed regions (7). Poverty had been increasing globally before the COVID-19 pandemic and 768 million people were hungry in 2020 (8). The African continent contributed more than one-third (282 million) of the hungry (8). It has been projected that hunger will increase globally due to the COVID-19 pandemic and even more in SSA (8). Additionally, further increases in child stunting and wasting in SSA are anticipated (9).

Further, the impact of COVID-19 on economic growth and food security in SSA remains severe (8). SSA economies are expected to experience recession and to continue to experience slower economic growth, with more disruptions of agriculture and production and trade-related constraints if efforts to control COVID-19 remain limited (10). This and resulting trade and fiscal deficits are likely to further adversely affect health and nutrition (11).

Countries and communities in SSA will also likely continue to face COVID-19 related disruptions to economic activity, livelihoods, and access to key nutrition and food security services (12). The continent has already reported an increase in prices for many staple crops (14), contributing to food insecurity. In our previous work in Burkina Faso, Ethiopia and Nigeria, we found evidence of increasing prices for key food items early during the COVID-19 pandemic, which may have contributed to lower dietary intake (13). In addition, we found that higher pulse prices during the COVID-19 pandemic were associated with the consumption of less diverse diets in the study countries (13). The potential impacts of COVID-19 can range from impacts on food availability, quality and prices, and impacts on vendors, markets and regulation during this time of the pandemic. On the other hand, restricted geographical access, changes in affordability due to the influence of global and local markets, desirability and convenience, and other factors could influence the food environment during the COVID-19 pandemic (14). Therefore, it is important that as the COVID-19 pandemic persists we continue to assess its impact on the nutrition and health of the households in SSA (12).

In this study, we investigated the continued impacts of COVID-19 on food consumption, food prices, dietary diversity, and quality using data collected during two rounds in five sub-Saharan African countries, Burkina Faso, Ethiopia, Ghana, Nigeria and Tanzania. We employed cross-sectional analysis using data collected from both rounds of the African Research, Implementation Science and Education (ARISE) Network COVID-19 surveys.

## Material and Methods

### Study setting

This survey was a repeated cross-sectional study with two rounds of data collection. The first round of the survey took place between July and November 2020. The study included six study sites from three countries, namely Nouna and Ouagadougou in Burkina Faso, Kersa and Addis Ababa in Ethiopia, and Ibadan and Lagos in Nigeria. In each country, one rural site and one urban site were selected. Rural sites included Nouna and Kersa, and a rural sub-area in Ibadan. The second round of the survey took place between July and December 2021 and involved all sites involved in round 1 and an additional three sites from two countries, namely the rural sites of Kintampo in Ghana, a rural subarea of Dodoma in Tanzania, and an urban area of Dar es Salaam in Tanzania. All nine sites from the five countries were included in the second round of data collection. The survey sites are shown in Figure 1.

**Figure 1:**
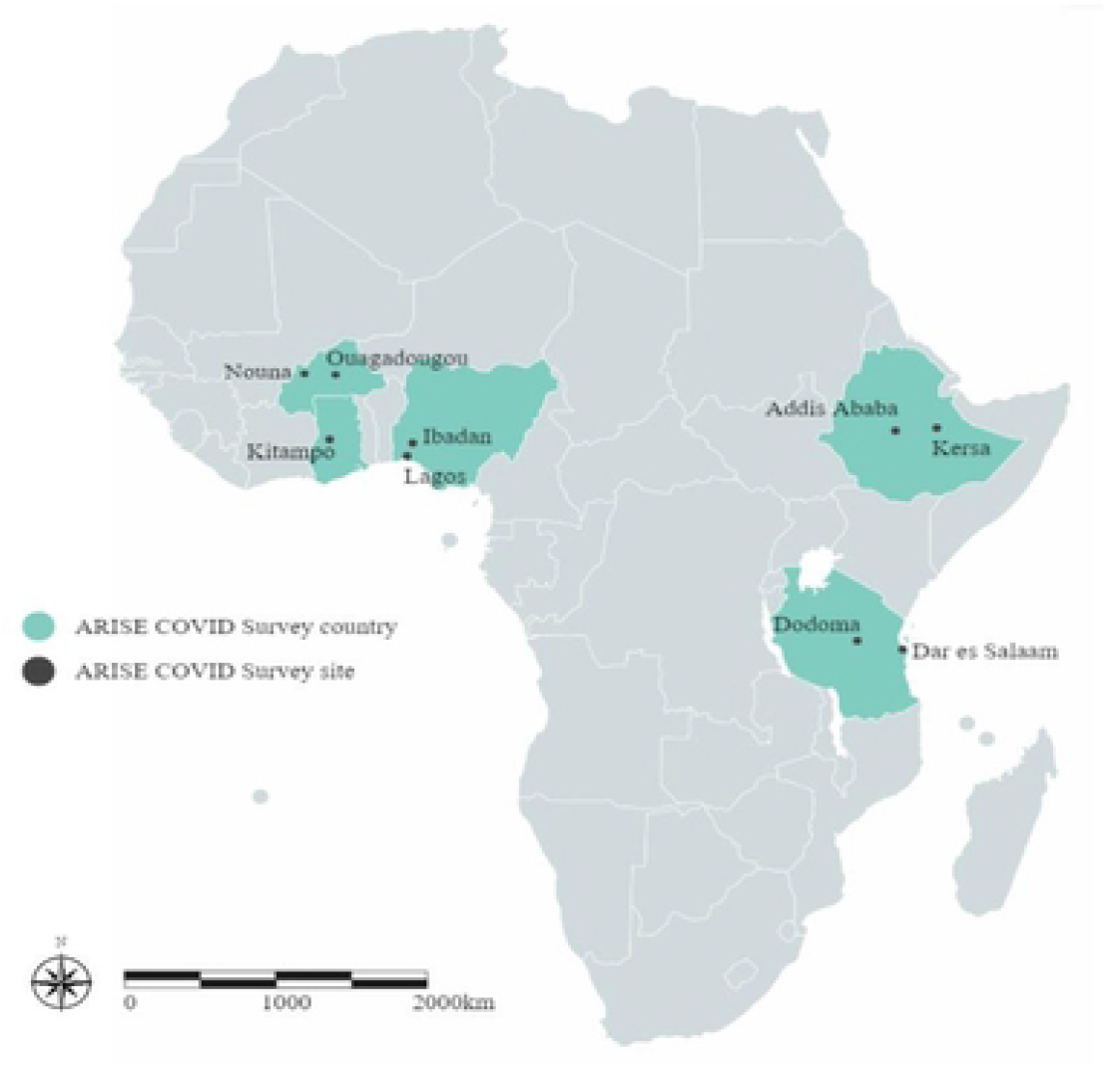
Map of ARISE sites for Round 2 of the COVID-19 studies.

The study countries and areas were selected based on existing collaborations and infrastructure available through the ARISE Network (15) and represent diverse settings across sub-Saharan Africa. More detailed information on the geographical and socio-demographic characteristics of the ARISE Network sites has been provided elsewhere (15, 16).

### Study design

The study used a mobile phone platform and computer-assisted telephone interviewing (CATI) to collect data. Research assistants obtained verbal informed consent before starting the interview. They conducted the interviews from study call centers.

Each country surveyed a minimum of 300 healthcare workers, 600 adults and 600 adolescents (except Ghana, where we surveyed 300 adults and 300 adolescents) in round 2. This analysis involved data from the adult surveys. We randomly selected a larger number of phone numbers of households from the sampling frame, to allow non-response, refusal or dropout. For the household surveys, 2,500 households were selected from each urban and rural site in Round 1, assuming that 60% of the households would respond to the survey. The assumption allowed us to reach the minimum number of 300 adults per site. From each household, an adult aged 20 years and above was selected for the interview.

The second round of data collection targeted the same adults previously surveyed in Round 1 (for Burkina Faso, Ethiopia and Nigeria) and additional new respondents to account for non-response and drop-outs. For new countries, the sample was obtained from existing sampling frames. The sampling frames were obtained from existing surveillance systems, the Health and Demographic Surveillance System (HDSS) in Burkina Faso, Ghana, Nigeria and rural Ethiopia (Kersa), and from a household survey in urban Ethiopia (Addis Ababa) that was established in the first round of data collection. In Tanzania, the sampling frames were obtained from the Dar es Salaam Urban Cohort Study (DUCS) and HDSS and the Dodoma HDSS.

The detailed design and field methods of the round 1 ARISE Network COVID-19 rapid monitoring survey have been published elsewhere (16). The design and methods of the round 2 survey are detailed on the Harvard University Center for African Studies website (https://africa.harvard.edu/files/africanstudies/files/arise_covid_survey_round_2_methods_brief_final.pdf).

Standardized tools used for the first survey round were updated for the second survey round. The local teams from each site reviewed the tools and updated the questions accordingly. Questionnaires were translated into the local languages. The sites recruited male and female research assistants who then received extensive training on data collection procedures, including the use of telephones and tablets to obtain verbal informed consent and input participant data into an electronic data collection system (Open Data Kit). Research assistants collected information on socio-demographic characteristics, including age, sex, head of household, household size, education, and occupation of respondents. Questions regarding food pricing, food security, and the dietary intake of respondents were also asked.

### Outcome Variables

The primary outcome of interest for this study was the Prime Diet Quality Score (PDQS). We also computed the diet diversity score (DDS) as a secondary outcome. The computation of the dietary indices, the PDQS, and DDS considered both Rounds of data collection. Briefly, respondents were asked about their frequency of consumption of 20 food groups commonly consumed in study areas over the preceding seven days.

### Prime diet quality score (PDQS)

We computed the PDQS, a measure of diet quality based on reported dietary intake. Previous studies have found associations between PDQS and birth outcomes, pregnancy-related morbidities, diabetes and cardiovascular disease (17-20). We classified foods into 20 food groups for the PDQS, including 14 healthy food groups (dark green leafy vegetables, other vitamin A-rich vegetables including carrots, cruciferous vegetables, other vegetables, whole citrus fruits, other fruits, fish, poultry, legumes, nuts, low-fat dairy, whole grains, eggs and liquid vegetable oils) and 6 unhealthy food groups (red meat, processed meats, refined grains and baked goods, sugar-sweetened beverages (SSBs), desserts and ice cream and fried foods obtained away from home and potatoes) based on criteria determined by previous studies (21, 22). Points were assigned for consumption of healthy food groups as: 0–1 serving/week (0 points), 2–3 servings/week (1 point), and ≥4 servings/week (2 points). Scoring for unhealthy food groups was assigned as: 0–1 serving/week (2 points), 2–3 servings/week (1 point) and ≥4 servings/week (0 points). Points for each food group were then summed to give an overall score (maximum score of 40).

### Dietary diversity score (DDS)

We also computed the Dietary diversity score (DDS) based on the classification suggested for the Food and Agriculture Organization (FAO)’s Minimum Dietary Diversity for Women (MDD-W) index. The MDD-W has been evaluated and considered a good measure of micronutrient adequacy among women (23, 24). We grouped the food consumed by study participants into 10 food groups as follows: 1) grains, white roots and tubers and plantains, 2) legumes (beans, peas and lentils), 3) nuts and seeds, 4) dairy, 5) meats, poultry and fish, 6) eggs, 7) vitamin A rich dark green vegetable, 8) other vitamin A-rich fruits and vegetables, 9) other vegetables and 10) other fruits. We divided the reported weekly consumption of the food groups by seven to obtain the daily frequency of consumption. If the food was eaten at least once daily during the previous week, it was considered to contribute to DDS, with a higher number indicating higher dietary diversity.

### Exposure Variables

We considered as exposures of interest changes in food pricing by comparing the time before and late during the COVID-19 pandemic for staples (maize, rice, cassava and teff), pulses (beans, lentils, peas, chickpeas), fruits (e.g. bananas, oranges, any locally available fruits), vegetables (e.g. spinach, cabbage, tomatoes, onions, any locally available vegetables) and animal source foods (e.g. beef, chicken, dairy, eggs, fish). We created a binary indicator indicating an increase compared to a decrease or no change in food prices (yes or no). The changes in food pricing were determined before and after March 2020 (first round), when the first case of COVID-19 was reported and also in the second round. Changes in food prices were self-reported by the participants.

We also considered food security status (e.g. went without eating for a whole day), and the impact of COVID-19 on income, employment and crop production. We considered respondent age (20-29, 30-39, ≥ 40 years), sex (female or male), education status (none or incomplete primary, primary school or incomplete secondary, secondary school or higher), head of household (yes or no), occupation (unemployed, farmer or casual labor, employed, student, self-employed or other), religion (none, Catholic, Muslim, Orthodox Christian, Protestant or other). Other exposures included household characteristics such as household size and wealth index score (tertiles). We computed the wealth index based on factor analysis on ownership of common household assets.

### Statistical Analysis

Descriptive statistics were used to describe numerical, tabular and graphical presentations of the data. We used means and standard deviations for continuous data and frequencies and percentages for categorical data to summarize social demographic characteristics, nutrition and food security across sites and rounds. We used Generalized Estimating Equation (GEE) linear regression models, to assess factors associated with the PDQS at the round 2 endpoint.

For those respondents with both round 1 and round 2 data, we evaluated the factors affecting diet quality in round 2 of the study. We conducted sensitivity analysis and assessed among respondents with both rounds 1 and 2 if adjusting for diet quality before the COVID-19 pandemic would change observed associations. We used GEE linear regression models, to assess factors associated with the PDQS at the round 2 endpoint accounting for diet quality at round 1.

Predictors of dietary quality were determined based on univariate selection. Variables associated with the outcomes at p<0.20 were included in the model. The final models include all selected covariates. Significance was determined at p<0.05. We used SAS version 9.4 for all analyses.

### Ethical approval and consent

The study obtained ethical approval from the Institutional Review Board at Harvard T.H. Chan School of Public Health, the Kintampo Health Research Centre Institutional Ethics Committee (Ghana), Nouna Health Research Center Ethical Committee and National Ethics Committee (Burkina Faso), the Institutional Ethical Review Board of Addis Continental Institute of Public Health (Ethiopia), University of Ibadan Research Ethics Committee and National Health Research Ethics Committee (Nigeria), and the Muhimbili University of Health and Allied Sciences and National Institute for Medical Research (Tanzania).

## Results

We analyzed data from 2,829 adults from the five countries. Overall, the study included 44% female respondents. Female participation was low in Nouna (15.4%), Ouagadougou (34.3%) and Kersa (14.8%). The mean (±SD) age of the respondents was 42.4 (±12.5) years. Most of the study respondents had no education or incomplete primary school education (37.9%), and 32.5% had secondary school education or higher. The mean (±SD) household size was 6 (±4) people. Most respondents were farmers or casual laborers (39.4%) and 32.5% were students or self-employed. A more detailed description of the demographic characteristics of the study individuals can be found in **Table 1**.

**Table 1.**
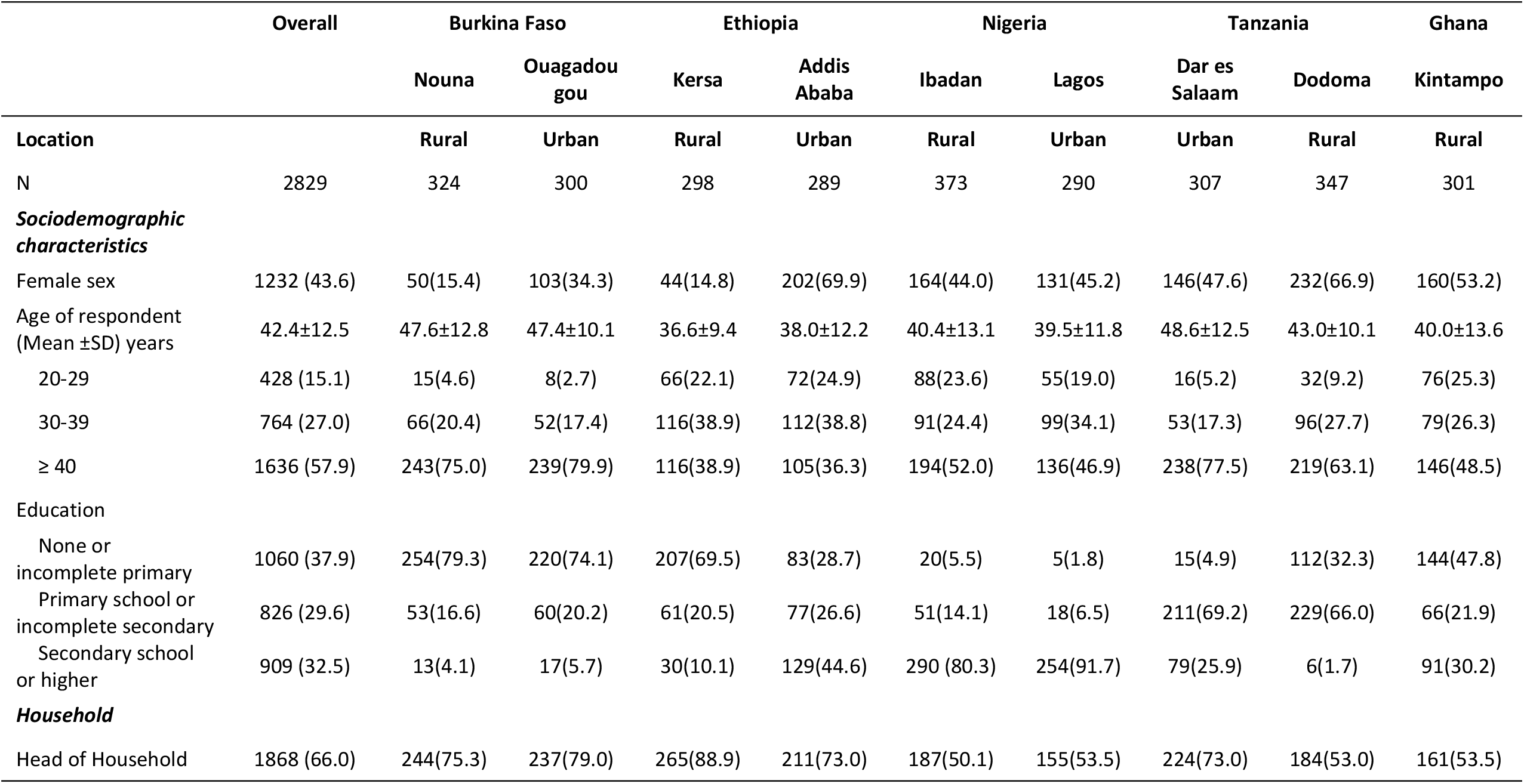
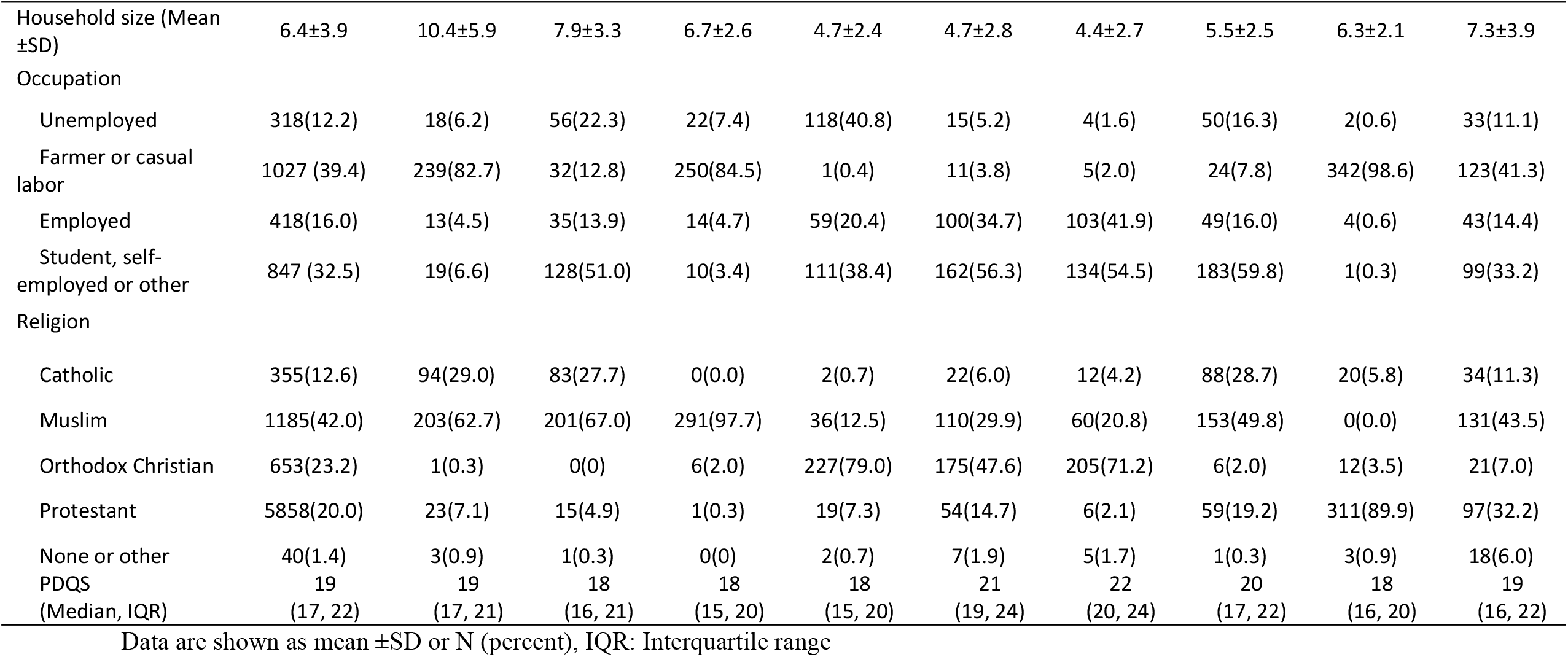
Demographic characteristics of the study individuals and households in Burkina Faso, Ethiopia, Nigeria, Tanzania and Ghana (N=2,829)

|  | Overall | Burkina Faso |  | Ethiopia |  | Nigeria |  | Tanzania |  | Ghana |
| --- | --- | --- | --- | --- | --- | --- | --- | --- | --- | --- |
|  |  | Nouna | Ouagadougou | Kersa | Addis Ababa | Ibadan | Lagos | Dar es Salaam | Dodoma | Kintampo |
| **Location** |  | **Rural** | **Urban** | **Rural** | **Urban** | **Rural** | **Urban** | **Urban** | **Rural** | **Rural** |
| N | 2829 | 324 | 300 | 298 | 289 | 373 | 290 | 307 | 347 | 301 |
| ***Sociodemographic characteristics*** |  |  |  |  |  |  |  |  |  |  |
| Female sex | 1232 (43.6) | 50(15.4) | 103(34.3) | 44(14.8) | 202(69.9) | 164(44.0) | 131(45.2) | 146(47.6) | 232(66.9) | 160(53.2) |
| Age of respondent (Mean ±SD) years | 42.4±12.5 | 47.6±12.8 | 47.4±10.1 | 36.6±9.4 | 38.0±12.2 | 40.4±13.1 | 39.5±11.8 | 48.6±12.5 | 43.0±10.1 | 40.0±13.6 |
| 20-29 | 428 (15.1) | 15(4.6) | 8(2.7) | 66(22.1) | 72(24.9) | 88(23.6) | 55(19.0) | 16(5.2) | 32(9.2) | 76(25.3) |
| 30-39 | 764 (27.0) | 66(20.4) | 52(17.4) | 116(38.9) | 112(38.8) | 91(24.4) | 99(34.1) | 53(17.3) | 96(27.7) | 79(26.3) |
| $\geq 40$ | 1636 (57.9) | 243(75.0) | 239(79.9) | 116(38.9) | 105(36.3) | 194(52.0) | 136(46.9) | 238(77.5) | 219(63.1) | 146(48.5) |
| **Education** |  |  |  |  |  |  |  |  |  |  |
| None or incomplete primary | 1060 (37.9) | 254(79.3) | 220(74.1) | 207(69.5) | 83(28.7) | 20(5.5) | 5(1.8) | 15(4.9) | 112(32.3) | 144(47.8) |
| Primary school or incomplete secondary | 826 (29.6) | 53(16.6) | 60(20.2) | 61(20.5) | 77(26.6) | 51(14.1) | 18(6.5) | 211(69.2) | 229(66.0) | 66(21.9) |
| Secondary school or higher | 909 (32.5) | 13(4.1) | 17(5.7) | 30(10.1) | 129(44.6) | 290 (80.3) | 254(91.7) | 79(25.9) | 6(1.7) | 91(30.2) |
| ***Household*** |  |  |  |  |  |  |  |  |  |  |
| Head of Household | 1868 (66.0) | 244(75.3) | 237(79.0) | 265(88.9) | 211(73.0) | 187(50.1) | 155(53.5) | 224(73.0) | 184(53.0) | 161(53.5) |
|  |  | Nouna | Ouagadougou | Kersa | Addis Ababa | Ibadan | Lagos | Dar es Salaam | Dodoma | Kintampo |
| Household size (Mean ±SD) | 6.4±3.9 | 10.4±5.9 | 7.9±3.3 | 6.7±2.6 | 4.7±2.4 | 4.7±2.8 | 4.4±2.7 | 5.5±2.5 | 6.3±2.1 | 7.3±3.9 |
| Occupation |  |  |  |  |  |  |  |  |  |  |
| Unemployed | 318(12.2) | 18(6.2) | 56(22.3) | 22(7.4) | 118(40.8) | 15(5.2) | 4(1.6) | 50(16.3) | 2(0.6) | 33(11.1) |
| Farmer or casual labor | 1027 (39.4) | 239(82.7) | 32(12.8) | 250(84.5) | 1(0.4) | 11(3.8) | 5(2.0) | 24(7.8) | 342(98.6) | 123(41.3) |
| Employed | 418(16.0) | 13(4.5) | 35(13.9) | 14(4.7) | 59(20.4) | 100(34.7) | 103(41.9) | 49(16.0) | 4(0.6) | 43(14.4) |
| Student, self-employed or other | 847 (32.5) | 19(6.6) | 128(51.0) | 10(3.4) | 111(38.4) | 162(56.3) | 134(54.5) | 183(59.8) | 1(0.3) | 99(33.2) |
| Religion |  |  |  |  |  |  |  |  |  |  |
| Catholic | 355(12.6) | 94(29.0) | 83(27.7) | 0(0.0) | 2(0.7) | 22(6.0) | 12(4.2) | 88(28.7) | 20(5.8) | 34(11.3) |
| Muslim | 1185(42.0) | 203(62.7) | 201(67.0) | 291(97.7) | 36(12.5) | 110(29.9) | 60(20.8) | 153(49.8) | 0(0.0) | 131(43.5) |
| Orthodox Christian | 653(23.2) | 1(0.3) | 0(0) | 6(2.0) | 227(79.0) | 175(47.6) | 205(71.2) | 6(2.0) | 12(3.5) | 21(7.0) |
| Protestant | 5858(20.0) | 23(7.1) | 15(4.9) | 1(0.3) | 19(7.3) | 54(14.7) | 6(2.1) | 59(19.2) | 311(89.9) | 97(32.2) |
| None or other | 40(1.4) | 3(0.9) | 1(0.3) | 0(0) | 2(0.7) | 7(1.9) | 5(1.7) | 1(0.3) | 3(0.9) | 18(6.0) |
| PDQS | 19 | 19 | 18 | 18 | 18 | 21 | 22 | 20 | 18 | 19 |
| (Median, IQR) | (17, 22) | (17, 21) | (16, 21) | (15, 20) | (15, 20) | (19, 24) | (20, 24) | (17, 22) | (16, 20) | (16, 22) |

Data are shown as mean ± SD or N (percent), IQR: Interquartile range

### Changes in income, employment, crop production and food security status

**Table 2** shows the impact of COVID-19 on income, employment, crop production and food security in the round 2 survey. Most respondents (49%) reported no change in income, and 37% reported loss or reduced income from farming, entrepreneurial activities, or business activities during the COVID-19 pandemic. Reported income losses from agriculture, entrepreneurial activities, or formal or informal business were highest in Kintampo (58%), Dar es Salaam (50%), and Ibadan (42%). Loss of or reduced salaries were reported most in Ouagadougou (37%) and Lagos (26%). The majority of respondents, however, reported that COVID-19 had not affected their employment status (only 7% reported lost employment). Loss of employment was relatively higher in Addis Ababa (16%), Ouagadougou (14%) and Ibadan and Nouna (at least 10%). Approximately 19% of all respondents across all sites reported lower agriculture production during the COVID-19 pandemic, with the highest declines reported in the rural sites of Kersa (64%), Nouna (37%) and Kintampo (27%). Food insecurity also affected respondents, with 45% reporting that they worried about food, and 30% said they had skipped a meal. Almost 10% had gone for an entire day without eating. The number of people reporting skipping a meal was highest in Ibadan (57%), Lagos (52%) and Kintampo (46%). Going for an entire day without eating was reported in Lagos (20%) and Ouagadougou (17%). Finally, social protection was limited, with less than 1% of the study sample reporting access to food aid, and 2% reporting access to cash transfers (not shown).

**Table 2.** Impact of COVID-19 on income, employment, crop production, and food security across 5 countries.

|  | Overall | Burkina<br>Nouna | Burkina<br>Ouaga | Ethiopia<br>Addis | Ethiopia<br>Kersa | Nigeri<br>a<br>Ibadan | Nigeri<br>a<br>Lagos | Tanzani<br>a Dar es<br>Salaam | Tanzani<br>a<br>Dodoma | Ghana<br>Kintamp<br>o |
| --- | --- | --- | --- | --- | --- | --- | --- | --- | --- | --- |
| **Impact of COVID-19 on income** |  |  |  |  |  |  |  |  |  |  |
| Income is unchanged | 49.4 | 61.6 | 29.7 | 48.1 | 82.1 | 43.3 | 37.7 | 40.9 | 78.1 | 34.8 |
| Lost/reduced salary | 12.1 | 4.3 | 36.8 | 9.1 | 0.7 | 11.9 | 26.1 | 7.9 | 0.6 | 6.4 |
| Lost/reduced income | 36.8 | 34.1 | 33.5 | 36.6 | 16.9 | 41.9 | 33.8 | 49.6 | 21.3 | 57.9 |
| Increased salary | 1.7 | 0.0 | 0.0 | 6.3 | 0.3 | 2.8 | 2.5 | 1.7 | 0.0 | 1.0 |
| **COVID-19 impact on employment** |  |  |  |  |  |  |  |  |  |  |
| None, unemployed | 31.1 | 46.8 | 30.0 | 43.9 | 10.4 | 16.5 | 13.6 | 41.0 | 55.9 | 20.7 |
| None, no change in employment status | 52.6 | 42.4 | 44.6 | 35.0 | 83.6 | 62.3 | 71.0 | 21.5 | 39.7 | 68.9 |

|  | Overall | Burkina<br>Nouna | Burkina<br>Ouaga | Ethiopia<br>Addis | Ethiopia<br>Kersa | Nigeri<br>a<br>Ibadan | Nigeri<br>a<br>Lagos | Tanzani<br>a Dar es<br>Salaam | Tanzani<br>a<br>Dodoma | Ghana<br>Kintampo |
| --- | --- | --- | --- | --- | --- | --- | --- | --- | --- | --- |
| Lost employment | 7.1 | 10.2 | 13.5 | 15.9 | 0.7 | 10.6 | 3.9 | 3.9 | 1.2 | 4.4 |
| Changed occupation | 3.1 | 0.3 | 2.3 | 4.8 | 1.7 | 3.3 | 5.2 | 4.3 | 1.5 | 5.0 |
| Other (specify) | 6.2 | 0.3 | 9.7 | 0.4 | 3.7 | 7.3 | 6.3 | 29.3 | 1.7 | 1.0 |
| **Crop production affected by COVID-19** |  |  |  |  |  |  |  |  |  |  |
| No | 36.6 | 44.3 | 23.7 | 4.2 | 32.7 | 6.5 | 31.4 | 67.4 | 91.9 | 22.2 |
| Yes, production has decreased | 18.6 | 36.6 | 9.4 | 1.0 | 63.6 | 11.1 | 7.7 | 4.6 | 8.1 | 26.9 |
| Yes, production has increased | 1.8 | 1.0 | 3.2 | 1.4 | 0.3 | 3.5 | 1.4 | 0.7 | 0.0 | 4.7 |
| No, I don't grow any crops | 43.0 | 18.2 | 63.7 | 93.4 | 3.4 | 78.9 | 59.6 | 27.4 | 0.0 | 46.3 |
| **Food insecurity** |  |  |  |  |  |  |  |  |  |  |
| Worried about food | 45.4 | 22.5 | 47.0 | 50.2 | 82.6 | 59.3 | 52.3 | 21.5 | 19.7 | 57.7 |
| Skipped a meal | 30.3 | 2.5 | 33.6 | 28.4 | 21.5 | 56.7 | 55.6 | 14.7 | 14.1 | 45.5 |
| Went for entire day without eating | 9.8 | 2.2 | 17.0 | 11.8 | 4.7 | 10.5 | 20.3 | 2.9 | 8.7 | 12.0 |

Respondents in rural regions of all sites were less likely to report changes in income or loss of salary compared to those residing in urban areas. Rural respondents in Ethiopia and Tanzania were also less likely to report reduced income, however, in other sites there were no rural-urban differences or income reductions that affected a greater proportion of rural respondents. Across all sites except Ethiopia, rural respondents were more likely to report no change in employment status. In rural sites more people reported worrying about food (except in Dodoma), however, going for an entire day without eating was more frequently reported in urban sites (except in Tanzania).

Among respondents that were in rounds 1 and 2, reports of disruptions to agricultural production were more prevalent in round 1 as at least 44% indicated that crop production had decreased. Additionally, more respondents had worried about running out of food (57%) and fewer had skipped a meal (21%) in round 1 (results not shown).

### Changes in prices for key food groups

In round 2 of the study, respondents reported that for all food groups, prices reported later during the COVID-19 pandemic were higher than typical prices in the previous year (Fig 2). Higher than expected prices were noted for all food groups across all sites for at least 80% of respondents, except in Dar es Salaam and Dodoma.

**Fig 2.**
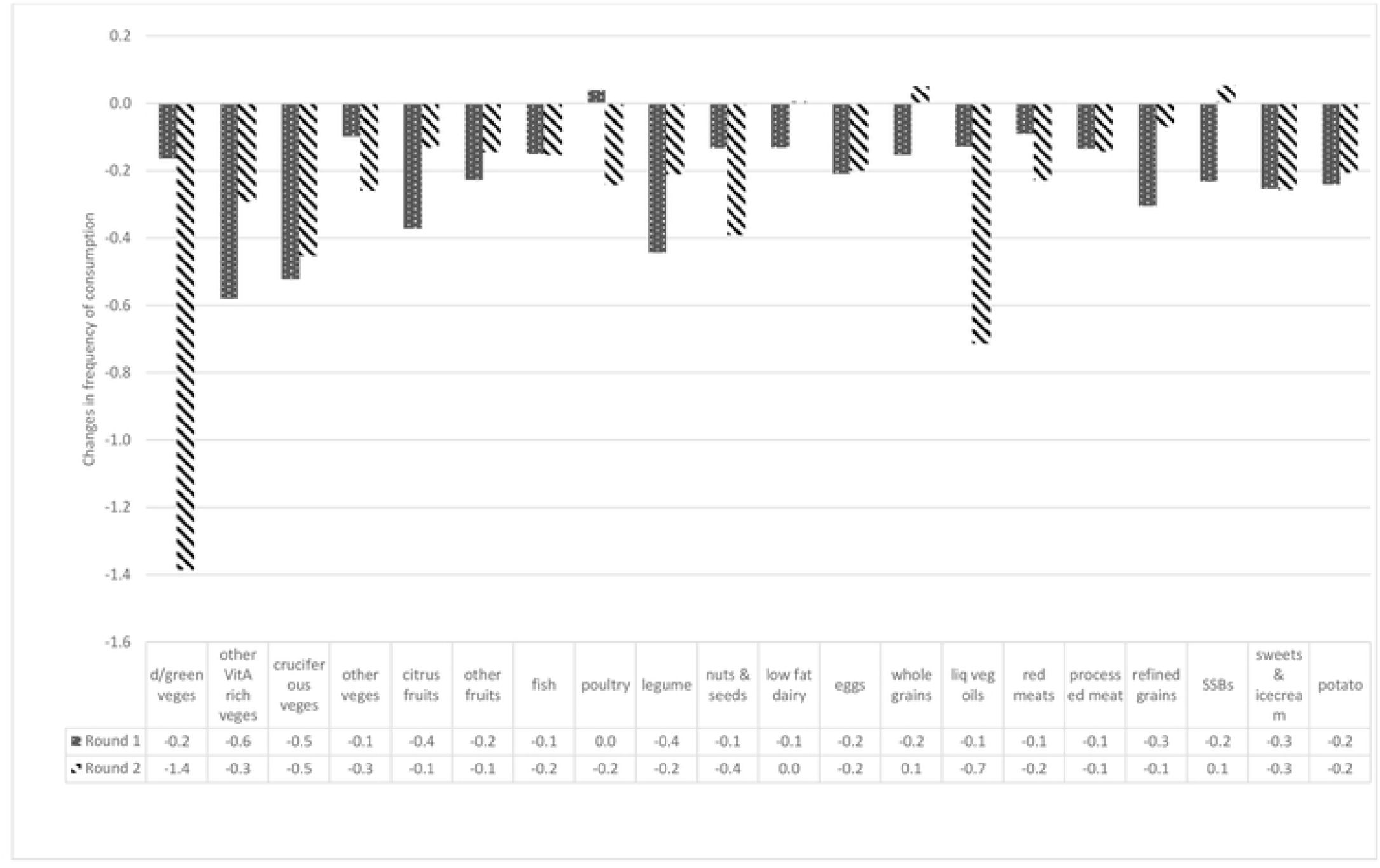
Changes in frequency of consumption of PDQS food groups comparing the time before the time of COVID-19 to the first and second round surveys.

Among respondents that were in rounds 1 and 2, at least 82% had reported increases in staple prices in round 1 compared to the time before the COVID-19 pandemic. For pulses, 77% reported increased prices, for fruits 66% reported higher prices, and 74% and 73% for vegetables and animal foods respectively. For all food groups except vegetables, higher prices were most frequently reported in round 1 (early during the COVID-19 pandemic) compared to round 2 (results not shown). The food groups where there were the largest differences in prevalence were fruits and animal-source foods.

### Food consumption and diet quality

#### Consumption of healthy foods

**Table 3** shows the frequency of consumption of healthy PDQS food groups across study sites in the five countries (Round 2). Consumption of dark green vegetables was high in Dar es Salaam, Kintampo and Dodoma, where 62%, 44%, and 38% of the respondents reported consuming these food groups four or more times a week, respectively. However, in Ethiopia, at least 69% reported consumption of dark green vegetables on one day or not at all in the previous week. Consumption of other vitamin A-rich vegetables was equally low in Burkina Faso, Ethiopia, Kintampo and Dodoma. Respondents reported low consumption of citrus and other fruits across most sites, except in Dar es Salaam, Ibadan, Lagos and Addis Ababa, where at least 22% of respondents reported citrus fruit consumption four or more times a week. In Addis Ababa, Kersa and Dodoma, most respondents reported consuming fish only once or not at all in the previous week. Poultry consumption was also low across the majority of sites, with more than 70% of respondents reporting consumption less than twice in the prior week, although intake was relatively higher in Lagos and Kintampo. In Addis Ababa (46%), Dar es Salaam (40%) and Dodoma (37%), respondents reported legume consumption most frequently (≥4 times a week). Whole-grain intake was higher in rural sites of Dodoma (55%), Kersa (52%), Kintampo (46%) and Nouna (42%). Consumption of nuts and seeds was low, as was dairy intake with at least 58% of the respondents reporting very low consumption in Ouagadougou, Addis Ababa, Kintampo and Tanzania. Egg consumption was also low in Tanzania, Burkina Faso and Ethiopia with 68% of the respondents reporting consumption less than twice in the previous week.

**Table 3.** Frequency of consumption of healthy PDQS food groups in Burkina Faso, Ethiopia, Nigeria, Tanzania and Ghana.

|  | Frequency (%) | Burkina<br>Nouna | Burkina<br>Ouaga | Ethiopia<br>Addis | Ethiopia<br>Kersa | Nigeria<br>Ibadan | Nigeria<br>Lagos | Tanzania<br>Dar es<br>Salaam | Tanzania<br>Dodoma | Ghana<br>Kintampo |
| --- | --- | --- | --- | --- | --- | --- | --- | --- | --- | --- |
| Dark green<br>vegetables | <=1 per week | 31.4 | 48.7 | 86.5 | 69.1 | 18.2 | 22.8 | 3.9 | 16.1 | 28.6 |
|  | 2-3 times per week | 52.2 | 43.7 | 11.1 | 24.2 | 55.2 | 50.0 | 33.9 | 45.8 | 27.6 |
|  | 4 or more times per week | 16.4 | 7.7 | 2.4 | 6.7 | 26.5 | 27.2 | 62.2 | 38.0 | 43.9 |
| Other Vit<br>A<br>vegetables | <=1 per week | 60.2 | 68.7 | 60.2 | 77.5 | 20.4 | 36.9 | 26.1 | 49.3 | 67.8 |
|  | 2-3 times per week | 37.0 | 25.7 | 29.8 | 20.5 | 58.7 | 42.8 | 45.9 | 31.7 | 20.9 |
|  | 4 or more times per week | 2.8 | 5.7 | 10.0 | 2.0 | 20.9 | 20.3 | 28.0 | 19.0 | 11.3 |
| Cruciferous<br>vegetables | <=1 per week | 45.4 | 41.3 | 63.7 | 40.9 | 48.8 | 43.8 | 48.2 | 62.0 | 71.8 |
|  | 2-3 times per week | 51.5 | 48.7 | 31.1 | 42.3 | 35.7 | 36.2 | 42.7 | 28.2 | 16.3 |
|  | 4 or more times per week | 3.1 | 10.0 | 5.2 | 16.8 | 15.6 | 20.0 | 9.1 | 9.8 | 12.0 |
| Other<br>vegetable | <=1 per week | 9.9 | 9.7 | 2.1 | 6.4 | 4.8 | 7.9 | 15.3 | 18.7 | 2.0 |
|  | 2-3 times per week | 49.1 | 39.3 | 2.1 | 14.4 | 30.6 | 30.3 | 30.6 | 38.3 | 6.3 |
|  | 4 or more times per week | 41.1 | 51.0 | 95.9 | 79.2 | 64.6 | 61.7 | 54.1 | 42.9 | 91.7 |
| Citrus | <=1 per week | 65.1 | 66.3 | 54.7 | 89.6 | 28.4 | 28.6 | 42.4 | 85.0 | 73.8 |
|  | 2-3 times per week | 32.4 | 26.0 | 22.8 | 9.4 | 46.1 | 44.1 | 35.5 | 11.2 | 14.6 |
|  | 4 or more times per week | 2.5 | 7.7 | 22.5 | 1.0 | 25.5 | 27.3 | 22.2 | 3.8 | 11.6 |
| Other fruit | <=1 per week | 57.5 | 69.7 | 65.4 | 89.9 | 30.6 | 34.1 | 22.5 | 74.6 | 70.8 |
|  | 2-3 times per week | 40.1 | 25.3 | 24.9 | 10.1 | 52.8 | 43.1 | 41.7 | 22.5 | 19.3 |
|  | 4 or more times per week | 2.5 | 5.0 | 9.7 | 0.0 | 16.6 | 22.8 | 35.8 | 2.9 | 10.0 |
| Fish | <=1 per week | 5.3 | 20.3 | 99.0 | 99.0 | 4.8 | 12.1 | 47.6 | 76.7 | 23.3 |
|  | 2-3 times per week | 56.2 | 44.7 | 1.0 | 0.7 | 31.1 | 25.2 | 39.4 | 17.8 | 18.3 |
|  | 4 or more times per week | 38.6 | 35.0 | 0.0 | 0.3 | 64.1 | 62.8 | 13.0 | 5.5 | 58.5 |
| Poultry | <=1 per week | 85.2 | 92.3 | 93.4 | 99.0 | 54.7 | 40.3 | 70.7 | 89.6 | 45.2 |
|  | 2-3 times per week | 14.5 | 7.3 | 4.8 | 1.0 | 31.6 | 30.0 | 26.7 | 8.9 | 20.6 |
|  | 4 or more times per week | 0.3 | 0.3 | 1.7 | 0.0 | 13.7 | 29.7 | 2.6 | 1.4 | 34.2 |
| Legumes | <=1 per week | 57.4 | 67.3 | 22.5 | 69.1 | 17.7 | 20.7 | 13.7 | 29.7 | 51.2 |
|  | 2-3 times per week | 41.1 | 26.0 | 31.5 | 22.8 | 62.2 | 50.0 | 45.9 | 33.4 | 28.6 |
|  | 4 or more times per week | 1.5 | 6.7 | 46.0 | 8.1 | 20.1 | 29.3 | 40.4 | 36.9 | 20.3 |
| Nuts and<br>seeds | <=1 per week | 26.9 | 63.0 | 89.6 | 72.2 | 32.4 | 31.4 | 49.2 | 77.8 | 27.9 |
|  | 2-3 times per week | 64.5 | 24.7 | 7.6 | 15.8 | 57.4 | 44.8 | 21.5 | 19.3 | 30.9 |
|  | 4 or more times per week | 8.6 | 12.3 | 2.8 | 12.1 | 10.2 | 23.8 | 29.3 | 2.9 | 41.2 |
| Dairy | <=1 per week | 38.0 | 76.7 | 58.8 | 38.9 | 17.7 | 22.4 | 59.3 | 74.1 | 61.1 |
|  | 2-3 times per week | 53.4 | 18.7 | 20.8 | 29.2 | 60.6 | 43.8 | 21.5 | 15.3 | 18.9 |
|  | 4 or more times per week | 8.6 | 4.7 | 20.4 | 31.9 | 21.7 | 33.8 | 19.2 | 10.7 | 19.9 |
| Eggs | <=1 per week | 68.2 | 73.7 | 73.7 | 74.5 | 16.6 | 19.0 | 73.0 | 87.6 | 52.8 |
|  | 2-3 times per week | 27.2 | 23.7 | 19.4 | 22.5 | 49.9 | 34.1 | 18.6 | 9.5 | 24.3 |
|  | 4 or more times per week | 4.6 | 2.7 | 6.9 | 3.0 | 33.5 | 46.9 | 8.5 | 2.9 | 22.9 |
| Whole<br>grains | <=1 per week | 16.4 | 41.3 | 66.4 | 12.4 | 26.8 | 35.5 | 44.0 | 34.6 | 31.9 |
|  | 2-3 times per week | 41.3 | 37.0 | 19.4 | 35.9 | 53.6 | 49.7 | 28.3 | 10.4 | 22.3 |
|  | 4 or more times per week | 42.3 | 21.7 | 14.2 | 51.7 | 19.6 | 14.8 | 27.7 | 55.0 | 45.9 |
| Liquid<br>vegetable<br>oils | <=1 per week | 16.4 | 42.0 | 19.0 | 17.5 | 4.6 | 9.3 | 57.0 | 9.8 | 31.2 |
|  | 2-3 times per week | 56.2 | 28.7 | 0.7 | 15.8 | 23.1 | 22.8 | 7.8 | 25.7 | 31.6 |
|  | 4 or more times per week | 27.5 | 29.3 | 80.3 | 66.8 | 72.4 | 67.9 | 35.2 | 64.6 | 37.2 |

\* Ouga - Ouagadougou; Burkina - Burkina Faso; Addis – Addis Ababa.

#### Consumption of unhealthy foods

**Table 4** describes the frequency of consumption of unhealthy foods. In Kintampo (36%), Ibadan (29%) and Lagos (27%) respondents reported higher red meat consumption (four or more times a week). Respondents in urban sites (Addis Ababa 22%, Lagos 22% and Dar es Salaam 18%) had relatively higher consumption of sugar-sweetened beverages, compared to rural sites. Processed meat was not commonly consumed, and sweets intake was low across all sites except Nigeria and Dar es Salaam. Finally, intake of refined grains was frequent across all study sites, and the majority of respondents in Kintampo (67%) reported consuming potatoes, roots and tubers four or more times a week.

**Table 4.** Frequency of consumption of unhealthy PDQS food groups in Burkina Faso, Ethiopia, Nigeria, Tanzania and Ghana.

|  | Frequency (%) | Burkina<br>Nouna | Burkina<br>Ouaga | Ethiopia<br>Addis | Ethiopia<br>Kersa | Nigeria<br>Ibadan | Nigeria<br>Lagos | Tanzania<br>Dar es<br>Salaam | Tanzania<br>Dodoma | Ghana<br>Kintampo |
| --- | --- | --- | --- | --- | --- | --- | --- | --- | --- | --- |
| Red meat | 4 or more times per week | 6.2 | 6.3 | 11.8 | 0.0 | 29.0 | 27.3 | 6.8 | 8.9 | 35.6 |
|  | 2-3 times per week | 59.6 | 24.3 | 25.6 | 0.7 | 44.2 | 46.2 | 34.9 | 26.5 | 23.3 |
|  | <=1 per week | 34.5 | 69.3 | 62.6 | 99.3 | 26.8 | 26.6 | 58.3 | 64.6 | 41.2 |
| Processed<br>meats | 4 or more times per week | 0.0 | 0.3 | 0.0 | 0.0 | 3.8 | 6.9 | 0.3 | 0.0 | 4.0 |
|  | 2-3 times per week | 5.9 | 0.3 | 0.0 | 0.0 | 27.4 | 29.0 | 4.2 | 0.0 | 9.6 |
|  | <=1 per week | 94.1 | 99.3 | 100.0 | 100.0 | 68.9 | 64.1 | 95.4 | 100.0 | 86.4 |
| Refined<br>grains | 4 or more times per week | 20.7 | 30.7 | 39.1 | 46.0 | 53.4 | 54.8 | 52.8 | 31.4 | 59.8 |
|  | 2-3 times per week | 49.1 | 42.0 | 29.8 | 41.3 | 38.9 | 36.2 | 28.3 | 21.9 | 21.9 |
|  | <=1 per week | 30.3 | 27.3 | 31.1 | 12.8 | 7.8 | 9.0 | 18.9 | 46.7 | 18.3 |
| SSBs | 4 or more times per week | 2.8 | 2.0 | 21.5 | 2.4 | 16.9 | 22.4 | 18.2 | 5.2 | 12.6 |
|  | 2-3 times per week | 42.0 | 20.3 | 13.8 | 22.2 | 46.1 | 36.2 | 29.6 | 12.7 | 21.9 |
|  | <=1 per week | 55.3 | 77.7 | 64.7 | 75.5 | 37.0 | 41.4 | 52.1 | 82.1 | 65.5 |
| Sweets<br>and ice<br>cream | 4 or more times per week | 8.6 | 5.0 | 3.5 | 3.4 | 12.1 | 12.4 | 3.3 | 14.1 | 6.3 |
|  | 2-3 times per week | 38.6 | 16.3 | 6.2 | 11.4 | 41.8 | 32.1 | 35.8 | 23.1 | 11.6 |
|  | <=1 per week | 52.8 | 78.7 | 90.3 | 85.2 | 46.1 | 55.5 | 60.9 | 62.8 | 82.1 |
| Potatoes,<br>roots and<br>tubers | 4 or more times per week | 1.9 | 0.3 | 10.7 | 18.8 | 23.9 | 29.0 | 37.1 | 14.4 | 66.5 |
|  | 2-3 times per week | 29.6 | 14.0 | 35.6 | 14.4 | 60.3 | 46.2 | 34.9 | 43.8 | 17.3 |
|  | <=1 per week | 68.5 | 85.7 | 53.6 | 66.8 | 15.8 | 24.8 | 28.0 | 41.8 | 16.3 |

\*SSBs - Sugar Sweetened Beverages; Ouga - Ouagadougou; Burkina - Burkina Faso; Addis – Addis Ababa.

#### Changes in consumption and diet quality

**Fig 2** shows changes in the frequency of consumption of PDQS food groups comparing the time before the COVID-19 pandemic to the first (July to November 2020) and second (July to December 2021) round surveys. The frequency of consumption of both healthy and unhealthy PDQS food groups was lower in Round 1 and Round 2 surveys (during the COVID-19 pandemic), compared to the period before the pandemic (except for SSB). In general, consumption declined most during the Round 1 survey compared with the period before COVID-19 and started to improve in Round 2. Data were captured around the same months for both surveys, which may decrease the influence of seasonality. However, there were exceptions. The frequency of consumption of dark green vegetables declined by 1 day and liquid vegetable oil consumption by 0.5 days on average in Round 2 compared to Round 1. Other food groups for which consumption was lower in Round 2 include poultry, nuts, and seeds.

#### Overall diet quality

The median PDQS (IQR) was 19 (17,22), and PDQS was highest in Nigerian sites and lowest in Ouagadougou, Addis Ababa, Kersa and Dodoma. **Fig 3** shows mean PDQS before COVID-19, early during the time of COVID-19 in 2020 (Round 1), and later during the COVID-19 pandemic in 2021 (Round 2). Overall, PDQS had returned to pre-COVID-19 levels across sites by Round 2 of the survey but remained low. There are, however, site-specific differences. In Nouna and Addis Ababa, PDQS was slightly higher in Round 2 of the survey compared to previous times. In all other sites, PDQS was lower in Round 2, compared to before the pandemic. Tanzania and Ghana did not participate in round 1 data collection.

**Fig 3.**
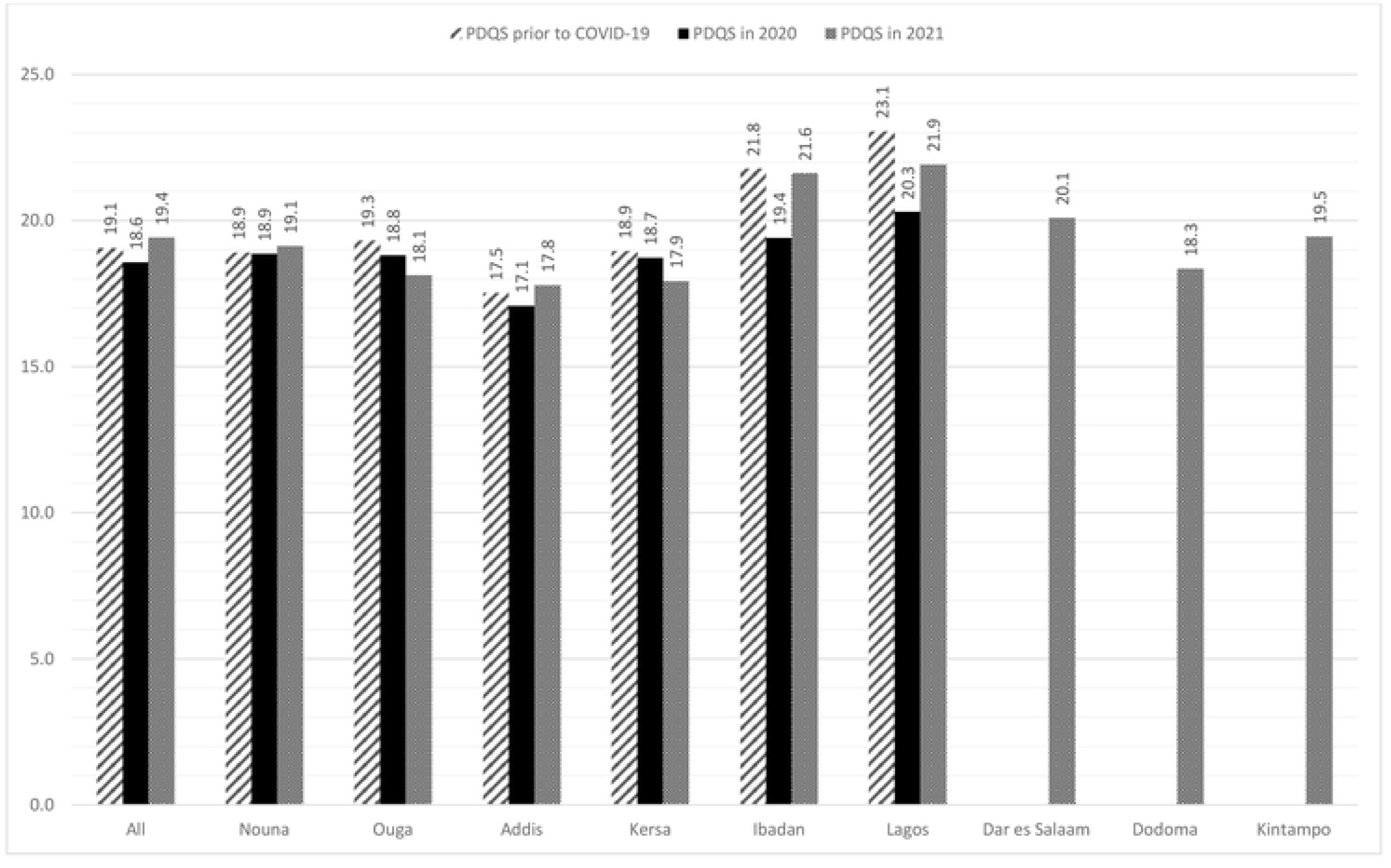
Mean Prime Diet Quality Scores (PDQS) before, early and later in the COVID-19 pandemic across five countries.

**Fig 4.**
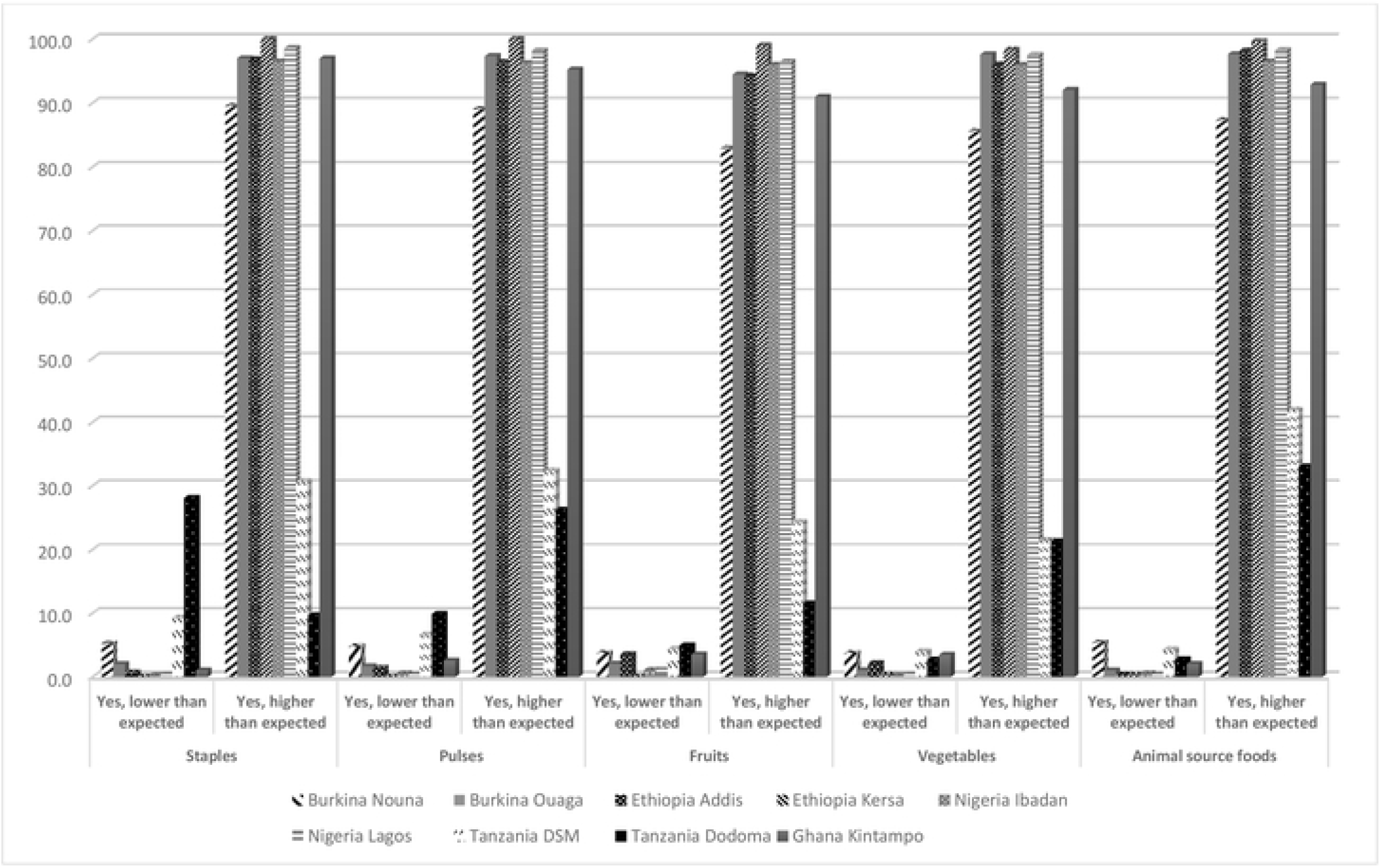
The proportion of people reporting changes in prices of key food groups during the COVID-19 pandemic, as compared to this time of the year in previous years.

**Table 5** shows the factors associated with diet quality across the five countries in Round 2 of the ARISE study. In a multivariate analysis, respondents residing in Nigeria (estimate: 2.84, 95% CI: 2.26,3.41) and Ghana (estimate: 0.93, 95% CI: 0.37,1.49) had higher PDQS compared to those residing in Burkina Faso. Respondents aged 30-39 years (estimate: 0.77, 95% CI: 0.35,1.19) and those 40 years or older (estimate: 0.72, 95% CI: 0.30,1.13) had higher PDQS compared to those aged 29 years or younger. Male respondents (estimate: -0.54, 95% CI: -0.88, -0.20) and those with no education had poorer diet quality (estimate: -0.40, 95% CI: -0.76, -0.03), and secondary school or higher education was associated with a higher PDQS (estimate: 0.73, 95% CI: 0.32,1.15) compared to having primary school education. Respondents who identified as Catholic (estimate: 0.66, 95% CI: 0.12,1.19) or Muslim (estimate: 0.51, 95% CI: 0.10,0.93) reported higher PDQS compared to Orthodox Christian respondents. Farmers and those engaged in casual labor reported lower PDQS (estimate: -0.60, 95% CI: -1.11, -0.09) compared to those that were formally employed.

**Table 5.** Factors associated with PDQS in Round 2 of the ARISE study across five countries.

|  | Univariate | Multivariate model<sup>a</sup> | Multivariate secondary analysis<sup>b</sup> |
| --- | --- | --- | --- |
|  | Estimate (95% CI) | Estimate (95% CI) | Estimate (95% CI) |
| PDQS before COVID-19 | 0.16(0.11,0.22)*** | - | 0.09(0.03,0.14)* |
| Country |  |  |  |
| Burkina Faso | ref | Ref | ref |
| Ethiopia | -0.78(-1.17,-0.39)*** | -0.37(-0.86,0.11) | -0.85(-1.54,-0.15)* |
| Nigeria | 3.11(2.72,3.49)*** | 2.84(2.26,3.41)*** | 1.93(0.82,3.04)** |
| Tanzania | 0.53 (0.14,0.91)* | -0.13(-0.74,0.49) | - |
| Ghana | 0.82(0.34,1.30)** | 0.93(0.37,1.49)** | - |
| Location |  |  |  |
| Rural area | -0.11(-0.39,0.11) | -0.13(-0.51,0.24) | 0.11(-0.66,0.88) |
| Urban | ref | ref | ref |
| Staple prices |  |  |  |
| No change or decreased | ref | ref | ref |
| Increased | 0.32(-0.01,0.65) | -0.01(-0.63,0.62) | 0.05(-1.28,1.37) |
| Pulse prices |  |  |  |
| No change or decreased | ref | ref | ref |
| Increased | 0.32(-0.01,0.66) | 0.21(-0.37,0.79) | 0.78(-0.37,1.94) |
| Fruits prices |  |  |  |
| No change or decreased | ref | ref | ref |
| Increased | 0.27(-0.04,0.58) | -0.45(-0.98,0.08) | -1.43(-2.33,-0.52) ** |
| Vegetables prices |  |  |  |
| No change or decreased | ref | ref | ref |
| Increased | 0.27(-0.03,0.60) | -0.15(-0.71,0.41) | 0.38(-0.64,1.39) |
| Animal source foods prices |  |  |  |
| No change or decreased | ref | ref | ref |
| Increased | 0.49(0.13,0.84)* | 0.41(-0.10,0.92) | -0.31(-1.42,0.81) |
| Age (years) |  |  |  |
| 20-29 | ref | ref | ref |
| 30-39 | 0.56(0.11,1.01)* | 0.77(0.35,1.19)*** | 0.35(-0.53,1.22) |
| ≥ 40 | 0.43(0.02,0.82)* | 0.72(0.30,1.13)** | -0.04(-0.91,0.84) |
| Sex |  |  |  |
| Female | ref | ref | ref |
| Male | -0.36(-0.64,-0.08)* | -0.54(-0.88,-0.20)** | 0.30(-0.36,0.96) |
| Education |  |  |  |
| None or incomplete primary | -0.65(-0.99,-0.33)*** | -0.40(-0.76,-0.03)* | -0.17(-0.77,0.43) |
| Primary school or incomplete secondary | ref | ref | ref |
| Secondary school or higher | 1.72(1.37,2.06)*** | 0.73(0.32,1.15)** | 1.25(0.47,2.03)* |
| Household head | -0.64(-0.93,-0.35)*** | 0.12(-0.24,0.47) | 0.08(-0.58,0.74) |
| Household size | -0.06(-0.10,-0.02)** | 0.02(-0.01,0.07) | 0.01(-0.04,0.07) |
| Religion |  |  |  |
| None | 0.01(-1.18,1.21) | -0.48(-0.64,1.61) | -2.52(-5.23,0.20) |
| Catholic | -0.56(-1.05,-0.08)* | 0.66(0.12,1.19)* | -0.79(-1.80,0.21) |
| Muslim | -0.89(-1.25,-0.54)*** | 0.51(0.10,0.93)* | -0.49(-1.30,0.33) |
| Orthodox Christian | ref | ref | ref |
| Protestant or other | -0.84(-1.26,-0.42)*** | 0.27(-0.25,0.78) | -0.59(-1.67,0.48) |
| Occupation |  |  |  |
| Unemployed | -2.20(-2.69,-1.70)*** | -0.39(-0.93,0.16) | 0.21(-0.69,1.12) |
| Farmer or casual labor | -2.06(-2.43,-1.70)*** | -0.60(-1.11,-0.09)* | 0.52(-0.35,1.39) |
| Employed | ref | ref | ref |
| Student, self-employed or other | -0.65(-1.03,-0.27)** | 0.01(-0.37,0.40) | 0.98(0.27,1.70)* |
| Effects of COVID-19 |  |  |  |
| None, unemployed | -0.94(-1.25,-0.62)*** | -0.38(-0.69,-0.06)* | -0.33(-0.90,0.23) |
| None, no change in employment status | ref | ref | ref |
| Lost employment | -0.01(-0.55,0.57) | 0.44(-0.09,0.98) | -0.31(-0.48,1.11) |
| Changed occupation | 0.18(-0.64,1.00) | 0.12(-0.64,0.88) | 0.20(-1.35,1.76) |
| Other (specify) | 0.82(0.23,1.42)* | 0.42(-0.15,1.00) | -0.58(-1.94,0.79) |
| COVID-19 Impact on income |  |  |  |
| Income is unchanged | ref | ref | ref |
| Lost or reduced salary from employer | 0.86(0.40,1.32)** | -0.19(-0.67,0.30) | -0.85(-1.62,-0.08)* |
| Lost or reduced income farming business | 0.37(0.06,0.68) | -0.13(-0.45,0.19) | -0.51(-1.07,0.05) |
| Increased salary from employer or farming business | 0.03(-1.11,1.17) | -0.36(-1.40,0.69) | -0.39(-2.16,1.37) |
| **Food security** |  |  |  |
| Went without eating for a whole day (past month) |  |  |  |
| No | ref | ref | ref |
| Yes | -0.71(-1.17,-0.24)** | -1.10(-1.53,-0.67)*** | -1.28(-2.03,-0.52)** |
| Own crop production affected |  |  |  |
| Unchanged | ref | ref | ref |
| Production has decreased | -1.06(-1.46,-0.67)*** | -0.87(-1.28,-0.46)*** | -1.04(-1.68,-0.41)** |
| Production has increased | 1.16(0.10,2.22)* | 0.18(-0.81,1.17) | 0.58(-1.13,2.29) |
| Does not farm | -0.22(-0.53,0.09) | -1.38(-1.74,-1.02)*** | -1.23(-1.84,-0.62)*** |
| Wealth index |  |  |  |
| Wealth tertile 1 | 0.02(-0.31,0.35) | -0.21(-0.51,0.10) | 0.14(-0.39,2.26) |
| Wealth tertile 2 | 1.07(0.71,1.42)*** | 0.48(0.14,0.81)* | 0.43(-0.16,1.02) |
| Wealth tertile 3 | ref | ref | ref |

Acronyms: PDQS: Prime Diet Quality Score, \* <0.05, \*\*<0.01, \*\*\*<0.001
a/Based on Round 2 data only
b/ Model restricted to individuals with round 1 and 2 data (N= 929) and adjusted for PDQS score prior to the COVID-19 pandemic (measured at Round 1).

Unemployed respondents had on average lower diet quality (estimate: -0.38, 95% CI: - 0.69, -0.06) compared to those whose employment status was unchanged during the COVID-19 pandemic. Those reporting going for an entire day without eating had lower PDQS than those who did not (estimate: -1.10, 95% CI: -1.53, -0.67). Lower crop production (estimate: -0.87, 95% CI: - 1.28, -0.46) or not engaging in farming (estimate: -1.38, 95% CI: -1.74, -1.02) were associated with lower PDQS compared to having unchanged levels of crop production. Being in the middle tertile of the wealth index (estimate: 0.48, 95% CI: 0.14,0.81) was associated with higher PDQS compared to being in the highest tertile.

In sensitivity analysis controlling for PDQS before the COVID-19 pandemic and restricted to the three countries with Round 1 data, PDQS before the COVID-19 pandemic was positively associated with PDQS later in the Round 2 survey (estimate 0.09, 95% CI: 0.03,0.14) after adjusting for other factors. We found country-level differences in PDQS, with respondents in Nigeria having higher PDQS (estimate 1.93, 95% CI: 0.82,3.04) and those in Ethiopia having lower PDQS (estimate: -0.85, 95% CI: -1.54, -0.15) compared to those residing in Burkina Faso. Higher fruit prices were associated with lower PDQS (estimate: -1.43, 95% CI: -2.33, -0.52). Secondary school education or higher was associated with higher diet quality (estimate: 1.25, 95% CI: 0.47,2.03) compared to primary school education, as was being self-employed or a student (estimates: 0.98, 95% CI:0.27,1.70) compared to being employed. Lower or reduced salary (estimate -0.85, 95% CI: -1.62, -0.08), lower crop production (estimate: -1.04, 95% CI: -1.68, - 0.41) and not farming (estimate -1.23, 95% CI: -1.84, -0.62) were associated with lower diet quality after adjusting for other factors.

## Discussion

We evaluated the impact of COVID-19 on food prices, food security, crop production, and diet quality, and the factors associated with diet quality two years into the COVID-19 pandemic in Burkina Faso, Ethiopia, Ghana, Nigeria and Tanzania. We found that food prices remained higher than expected during the pandemic, that consumption of healthy food groups remained low, and that diet quality showed signs of slight recovery since 2020 but remained poor. Food insecurity, low agriculture production, and occupation were negatively associated with PDQS, while higher socioeconomic status was positively associated with diet quality.

In this study, we found evidence of the continued impact of COVID-19 on food prices, with the majority of respondents indicating that food prices were higher than before the pandemic. In a study conducted across several countries early in the COVID-19 emergency in June 2020, there was evidence that the prices of maize, sorghum, imported rice and rice in SSA were higher than expected (25). Price increases were attributed to movement restrictions and lockdowns, with economic factors such as exchange rate, and inflation also contributing (25). Other studies indicated that a global slowdown due to COVID-19 and its related impact on Gross Domestic Product (GDP) could lead to impacts on food security and consumption, through increased unemployment, reduced trade and decreased production (26).

In this study, we found that lower agricultural production, not participating in crop production, as well as food insecurity, were associated with the consumption of lower-quality diets. The impacts of COVID-19 on agricultural production and food security have also been documented in previous studies. In one study, the COVID-19 pandemic was shown to impact bean production by small-scale farmers in SSA, with disruptions in access to seeds, inputs, and labor, among other things (27). Lower agricultural production can impact the consumption of quality diets by decreasing the availability of foods and increasing prices for nutritious foods. In Nigeria, disruptions due to COVID-19 restrictions early in the pandemic included lockdowns, curfews and travel restrictions in urban areas and these affected the ability of local authorities to support agricultural production, disrupted trade and decreased access to healthy diets (28). Although restrictions across countries have been lifted, the lasting impact may be due to impacts on regional and international food trade that have affected food prices (28).

The impacts of COVID-19 on the African continent, particularly on food security and diets, are expected to be dire, given that purchasing power and social safety nets are already limited (29). However, we found that diet quality and diversity scores showed small increases later during the COVID-19 crisis and were closer to pre-COVID states. However, despite these increments, it was still clear that consumption of key food groups such as dark green vegetables, other vegetables, poultry, nuts and seeds, and liquid vegetable oils were declining. The consumption of red meat also showed signs of declining. For the DDS food groups, the majority of respondents reported decreased consumption, with notable declines for dark green vegetables, other vegetables, staples, meats and fruits. Further, the noted recovery of diet quality scores is small and both diet quality and diversity remain low across all the countries assessed. Moreover, there were country-specific differences with greater recovery in diet quality experienced in sites in Nigeria and Addis Ababa. In contrast, other sites continued to experience declining diet quality and diversity. These differences could reflect the differences in the states’ capacity to mitigate against the COVID-19 impacts and the availability of social protection. In this study, however, the availability of social protection to address the impacts of COVID-19 was very limited.

In several countries, the impacts of COVID-19 on food prices and quality have been due to decreased economic activity, lower income and declines in employment and income. We found that loss of employment was infrequently reported. However, nearly 50% of the respondents indicated that they had lost income from employment, farming or business activities. The impacts of the loss of income from employment and income-generating activities on diets can be through loss of purchasing power (30). The effects of COVID-19 related income shocks on food security and diets have been reported in Uganda and Kenya, with poor households most affected (31, 32). For example, loss of income during COVID-19 was reported by farmers in Burundi (36%), Uganda (20%), and Kenya (3%). In our study, loss of income from farming and business was greatest in Kintampo, Dar es Salaam and Ibadan, with rural areas more likely to experience a loss due to impacts on agriculture. Previous studies have, however, indicated that rural areas and farming communities might be more resilient to the impacts of COVID-19 due to shorter value chains and reliance by smallholder farmers on their production (30). In our study, the impacts on rural and urban locations depend on the country context; for example, the largest declines in DDS were observed in urban sites such as Ouagadougou and rural sites of Kersa and Nouna.

Studies have also indicated that the greatest impact of COVID-19 in the African context has been felt by those with low socioeconomic status, informal workers, and those reliant on daily income (30). We found that the older or more educated respondents with higher socioeconomic status had higher diet quality in the study. We also found that those engaged in farming or casual labor in our study tended to have lower diet quality, reinforcing this observation. These individuals are less likely to have enough savings to cope with shocks such as the COVID-19 pandemic and may have been highly vulnerable even before the shock. Economic stimulus packages and investment in sectors such as agriculture have been proposed as possible solutions to address the challenges posed by the COVID-19 pandemic in Africa and to promote recovery (27). However, these were not readily available to communities and respondents in our study settings.

Our study has several strengths. First, we conducted a repeated cross-sectional study in 3 countries, allowing us to track changes in food security and diet intake at different times during the COVID-19 pandemic. Further, we collected data on the same individuals in some contexts. A limitation of the study is that we had a loss to follow-up in round 2 of the survey, limiting our ability to look at changes over time prospectively. Loss to follow-up is, however, expected in telephone surveys, especially in our study contexts. Also, possible measurement errors as a result of self-reported dietary intake and food pricing.

## Conclusion

We found continuing effects of COVID-19 on food prices, and low dietary intake and quality across the study sites. While there were improvements in diet quality later during the pandemic, consumption of healthy diets remained low, and improvements lagged across various sites. High food prices and poor diet quality experienced in the study have health implications. Efforts should continue to improve diet quality through mitigation measures, including social protection for people for sustained nutrition recovery.

## Data Availability

Data was collected by a consortium of partners. Data release for some countries may require authorization.

## Acknowledgments

We thank all study participants and data collectors for contributing to this study. The survey team in Ghana is grateful for support from the Kintampo Health Research Centre of Ghana Health Service, and the community leadership of Kintampo North Municipality and Kintampo South District. We acknowledge institutional support from Harvard T.H. Chan School of Public Health, Boston, MA; Harvard University Center for African Studies, Boston, MA; Heidelberg Institute of Global Health, Germany and the George Washington University Milken Institute of Public Health, Washington, DC.

## Abbreviations

ARISE: African Research, Implementation Science and Education Network
DDS: Dietary diversity score
DUCS: Dar es Salaam Urban Cohort Study
HDSS: Health and Demographic Surveillance System
MDD-W: Minimum Dietary Diversity for Women
PDQS: Prime Diet Quality Score
SSA: Sub-Saharan Africa
SSBs: sugar-sweetened beverages

## Authors’ contributions

AI and IM conceived and designed the study, analyzed the data, and drafted the manuscript. WWF was the principal investigator for the parent study, conceived the study, designed the study, interpreted the data, and guided revisions of the manuscript. YB designed the study, interpreted the data, and guided revisions of the manuscript. EAP, AT, DD, PZ, Mo, NA, AC, FW, FM, BL, EH, DW, SWA, KPA, TB, JK, AO, AS, AS, SV, and ES designed the study, interpreted the data, and guided revisions of the draft manuscript. All authors read and approved the final manuscript.

## Data sharing

Data described in the manuscript, code book, and analytic code will be made available upon request pending approval.

